# A cross-sectional study to assess beta cell function in individuals with recently diagnosed young onset type 2 diabetes mellitus and its complications

**DOI:** 10.1101/2022.08.22.22279100

**Authors:** Shamharini Nagaratnam, Subashini Rajoo, Mohamed Badrulnizam Long Bidin, Nur Shafini Che Rahim, Sangeetha Tharmathurai, Masita Arip, Yee Ming Ching, Siew Hui Foo

**Affiliations:** Endocrine Unit, Department of Medicine, Selayang Hospital, Selangor, Malaysia; Endocrine Unit. Department of Medicine, Kuala Lumpur Hospital, Kuala Lumpur, Malaysia; Department of Pathology, Kuala Lumpur Hospital, Kuala Lumpur, Malaysia; Department of Ophthalmology, Kuala Lumpur Hospital, Kuala Lumpur, Malaysia; Allergy and Immunology Centre, Institute for Medical Research, National Institute of Health, Selangor, Malaysia

## Abstract

The primary objective of this study was to assess beta cell function of recently diagnosed young onset type 2 diabetes mellitus (T2DM) individuals. The secondary objective examined the association between C-peptide with metabolic factors and diabetes complications. A cross-sectional study was conducted for young onset T2DM individuals aged 18-35 years with disease duration not more than 5 years. Plasma basal and stimulated C-peptide was measured before and after intravenous glucagon injection. Demographic data, medical history and complications were obtained from medical records and clinical assessment. A total of 113 participants with young onset T2DM with mean age of 29 years and median disease duration of 24 months were included in this study. The median (interquartile range) basal and stimulated C-peptide was 619 (655) pmol/L and 1231(1024) pmol/L. Adequate beta cell function was present in 78-86% of the participants based on the basal and stimulated C-peptide levels. Obesity and hypertension were independently associated with higher basal and stimulated C-peptide while diabetic kidney disease was independently associated with higher basal C-peptide. We found most recently diagnosed young onset T2DM have adequate beta cell function. Elevated C-peptide levels associated with obesity, hypertension and diabetic kidney disease suggests insulin resistance as the key driving factor for complications.

## Introduction

Young onset type 2 diabetes mellitus (T2DM) is defined as onset of T2DM before the age of 40 years in the absence of secondary causes (1). Studies have shown an alarming increase in the prevalence of young onset T2DM globally, more so in Asia in the last few decades. This subset of individuals has been associated with accelerated disease progression and premature complications (2). There is also higher rates of insulin commencement and intensification of treatment regime early in the disease compared to the usual onset (1). The pathophysiology of T2DM in the young has been thought to be similar to the usual onset with an interplay of beta cell dysfunction, insulin resistance and obesity related mechanisms (2,3). There is limited data with regards to beta cell function in the early stage of the disease in this population. This is a pilot study performed in the country to assess beta cell function of recently diagnosed young onset T2DM individuals using fasting and glucagon stimulated C-peptide levels. C-peptide produced in equal amounts to insulin is an objective marker of beta cell function. Glucagon stimulation test (GST) is an easily performed test to assess stimulated C-peptide with good sensitivity and reproducibility in clinical practice (4). The secondary objective of this study was to examine the associations between C-peptide levels with metabolic parameters and diabetes related complications.

## Material and Methods

This is a cross-sectional study involving young onset T2DM individuals attending diabetes clinic in two urban tertiary hospitals in Malaysia carried out between September 2019 to December 2020. Individuals aged between 18 to 35 years diagnosed with T2DM for not more than five years were recruited using universal sampling method. We excluded individuals with chronic kidney disease stage 3 and above, concurrent infection or inflammatory disease, recent diabetic ketoacidosis or hyperglycaemic hyperosmolar state in the last three months, positive diabetes autoantibodies, fasting capillary blood glucose less than 4.0 or more than 13.9 mmol/L on the day of testing and those who were pregnant (5). Written informed consent was obtained from all study participants prior to commencement of the study. This study was approved by the Medical Research and Ethics Committee (MREC) of Ministry of Health Malaysia.

Study participants were recruited during their routine clinical visit to diabetes clinic. Individuals with unknown diabetes mellitus autoantibodies status were screened prior to enrolment. This study involved only a single study visit. Study participants were required to fast overnight for at least eight hours and to omit all insulin and oral antidiabetic agents on the morning of testing. On the day of testing, anthropometric measurements (weight, height, waist circumference) were taken using calibrated tools. Vital signs and capillary blood glucose pre-procedure were checked. GST was performed if fasting capillary glucose fell between 4.0 to 13.9 mmol/L. Basal C-peptide and blood glucose levels were sampled prior to administration of intravenous glucagon 1 mg. After 6 minutes, stimulated blood glucose and C-peptide samples were collected. Study participants were monitored for adverse effects for 15 minutes post testing. Information regarding demography, disease history, co-morbidities, complications and treatment were gathered from medical records and clinical assessment. Macrovascular complication was defined as established history of ischemic heart disease, stroke or peripheral vascular disease. With regards to microvascular complications, retinopathy was assessed based on records of slit lamp examination or fundus camera performed in the last 12 months. Participants with no recent assessment were referred to ophthalmology for evaluation. Diabetic kidney disease (DKD) was assessed based on two or more urinary spot quantification of proteinuria or urine albumin creatinine ratio (ACR) performed in the last 12 months. Microalbuminuria and overt nephropathy were defined as urine ACR of 3-30 mg/mmol and >30 mg/mmol respectively (6). Peripheral neuropathy was diagnosed based on diminished sensation to standardised monofilament examination or reduced vibration on graduated tuning fork testing. Glycated haemoglobin (HbA1c) performed in the last three months was used as a measure of glycaemic control. C-peptide analysis was performed in a centralised laboratory using IMMULITE-2000 C-peptide (Siemens), a solid phase, two-site chemiluminescence immunometric assay with a coefficient of variation (CV) of less than 5%. Study participants were screened for diabetes autoantibodies including anti-islet cell antibodies (ICA), anti-glutamic acid (GAD) antibodies and anti-islet tyrosine phosphatase (IA2) antibodies using commercially available ELISA kits (Medipan GmbH, Germany).

All statistical analysis was performed with Statistical Package for Social Science (SPSS) Version 22.0. Most continuous data was found to be not normally distributed, therefore expressed as median and interquartile range (IQR). Categorical variables were described as frequency or percentage. Bivariate analysis using Mann Whitney U and Kruskal Wallis test were used to examine association between categorical data. Spearman’s rank correlation coefficient was used to assess relationship between continuous data. We proceeded with multivariate linear regression analysis using variables with p-values <0.25 from bivariate analysis and clinically important outcome variables based on biological plausibility. Stepwise linear regression was applied for variables selection. The final model was tested for autocorrelation (Durbin-Watson test), multicollinearity and homoscedasticity. The significance level for all analyses was p < 0.05.

## 3. Results

### Baseline characteristics (Table 1)

A total of 113 participants with young onset T2DM with median age of 29 years and median disease duration of two years were included.There was a female preponderance at 62% with the predominant ethnicity being Malay, at 71%.Based on the World Health Organization (WHO)’s body mass index (BMI) cut off for obesity in Asians of 27.5 kg/m^2^, 74% of the study population were obese (7). In addition, nearly 90% of them had abdominal obesity with a waist circumference of ≥ 90 cm in males or ≥80 cm in females (8). Almost all had concomitant dyslipidaemia while approximately one third had hypertension. A total of 89% of the study population fulfilled the International Diabetes Federation (IDF) criteria of metabolic syndrome (9). Majority, had a family history of T2DM involving at least one first degree relative while 14% were active smokers. The average glycaemic control of the population studied was poor, with a median (IQR) HbA1c of 8.5 (4.1) % (69 (21) mmol/mol). More than three quarters of the study population were initiated on insulin within six months from diagnosis, but only 18% were successfully weaned off insulin after a median disease duration of two years. There were no documented macrovascular complications. However, microvascular complications were present in 38% of the cohort. DKD was the most common microvascular complication at 35% in the form of microalbuminuria. Retinopathy and peripheral neuropathy were observed in less than 10% of the cohort.

**Table 1:**
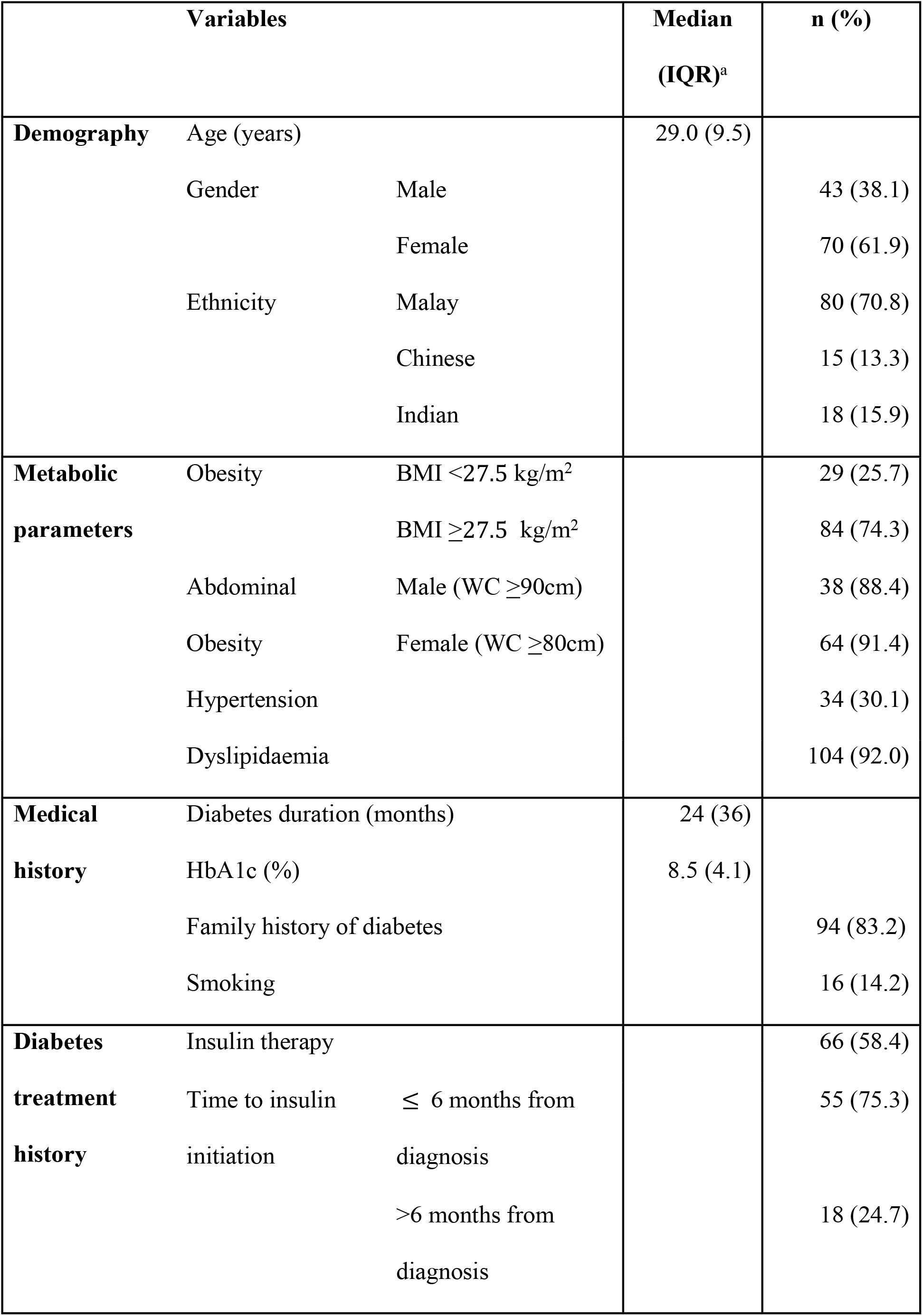
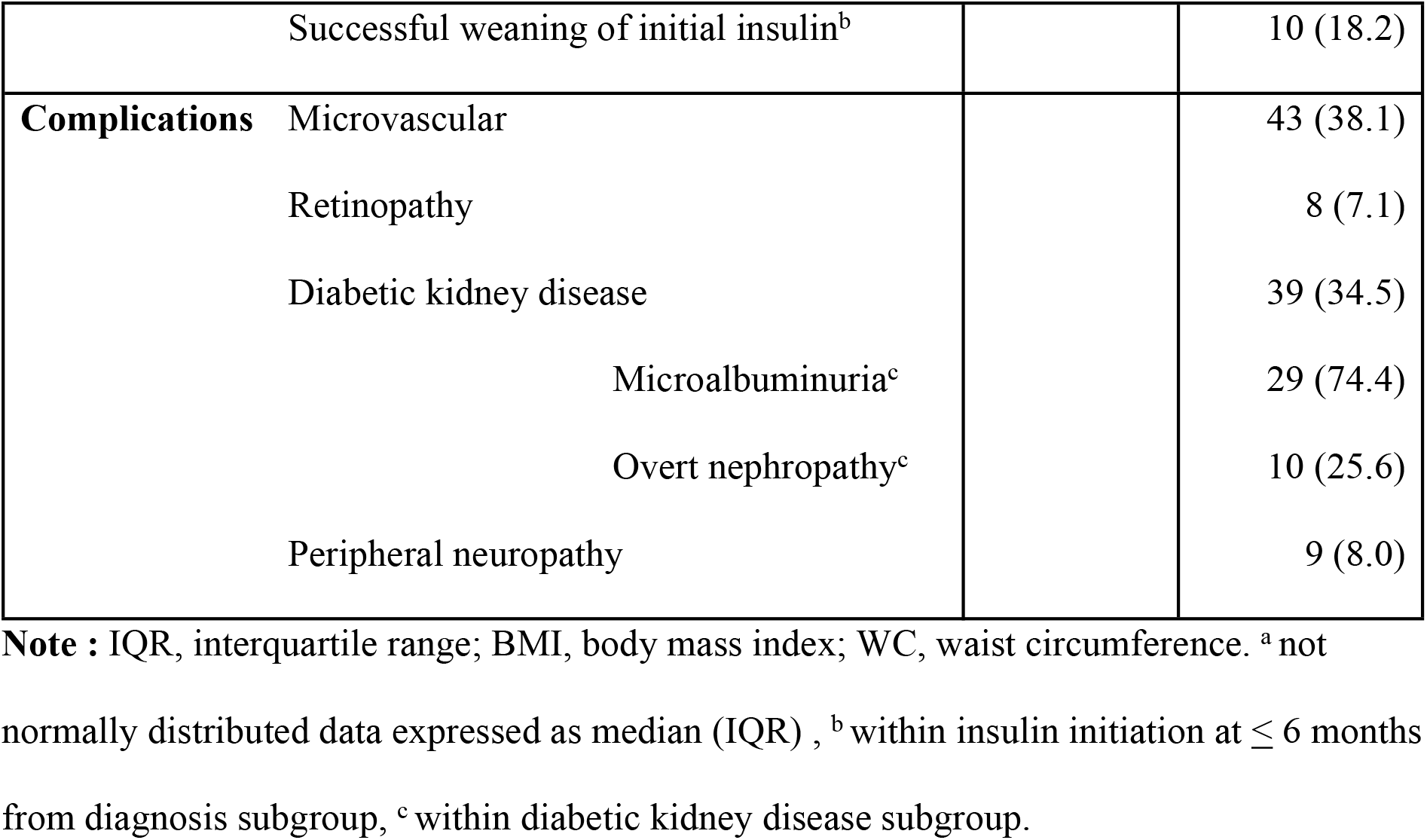
Baseline characteristics of the study population (N=113)

### Glucagon stimulation test results (Table 2)

The C-peptide levels had a median (IQR) basal value of 619 (655) pmol/L and 1231 (1024) pmol/L with stimulation. There was a strong positive correlation between fasting and stimulated C-peptide levels, rs=0.92, p <0.001. Adequacy of beta cell function was defined as basal C-peptide level of more than 250 pmol/L and stimulated C-peptide level of more than 600 pmol/L (10). Majority of the study participants had adequate beta cell function based on the basal and stimulated C-peptide levels at 86% and 78% respectively. Within the subgroup of participants on insulin as current therapy, there was still a high proportion registering adequate beta cell function at 77% and 70% respectively based on the basal and stimulated C-peptide levels. Transient and self-limiting nausea was reported in 10 participants immediately after the procedure.

**Table 2:** Glucagon stimulation test results.

| Variables |  | Median (IQR) | n (%) |
| --- | --- | --- | --- |
| **Glucose (mmol/L)** | Fasting glucose | 7.5 (3.3) |  |
|  | Stimulated glucose | 8.3 (3.7) |  |
| **C-peptide (pmol/L)** | Basal C-peptide | 619.0 (655.0) |  |
|  | Glucagon stimulated C-peptide | 1231.0 (1024.0) |  |
| **Beta cell function** | Adequate basal beta cell function <sup>a</sup> |  | 97.0 (85.8) |
|  | Adequate stimulated beta cell function <sup>b</sup> |  | 88.0 (77.9) |
|  | For participants on insulin as current therapy: |  |  |
|  | i. Adequate basal beta cell function<sup>a</sup> |  | 51.0 (77.0) |
|  | ii. Adequate stimulated beta cell function<sup>b</sup> |  | 46.0 (69.7) |

**Note:** Results expressed in median (IQR) for not normally distributed variables and percentage for categorical variables. <sup>a</sup> defined as C-peptide >250 pmol/L , <sup>b</sup> defined as C-peptide >600 pmol/L.(10)

### Factors associated with basal and stimulated C-peptide levels

Table 3 shows the bivariate analysis between basal and stimulated C-peptide levels with baseline clinical characteristics, metabolic parameters and diabetes related complications. Both basal and stimulated C-peptide were significantly associated with gender, disease duration, age of disease onset, HbA1c, insulin therapy, obesity, waist circumference and hypertension (p <0.05). In addition, stimulated C-peptide was also significantly associated with current age, smoking and dyslipidaemia (p <0.05). There was no significant association between basal and stimulated C-peptide with diabetes related complications. Multivariable linear regression analyses were performed to determine factors independently associated with basal and stimulated C-peptide levels (Table 4 and 5). We included 11 variables with p-values <0.25 from the bivariable analysis and clinically important outcome variables studied into the preliminary main effect model (current age, gender, smoking, age of disease onset, disease duration, HbA1c, obesity, insulin therapy, dyslipidaemia, hypertension and DKD) for both basal and stimulated C-peptide. Basal C-peptide was found to have independent association with insulin therapy, hypertension, obesity, gender, DKD and smoking (Table 4). Stimulated C-peptide was found to have independent association with insulin therapy, hypertension, obesity, gender, and duration of disease (Table 5). Among the metabolic parameters, both obesity and hypertension independently predicted a higher basal and stimulated C-peptide level. As for the diabetes related complications, DKD independently predicted a higher basal C-peptide level.

**Table 3:**
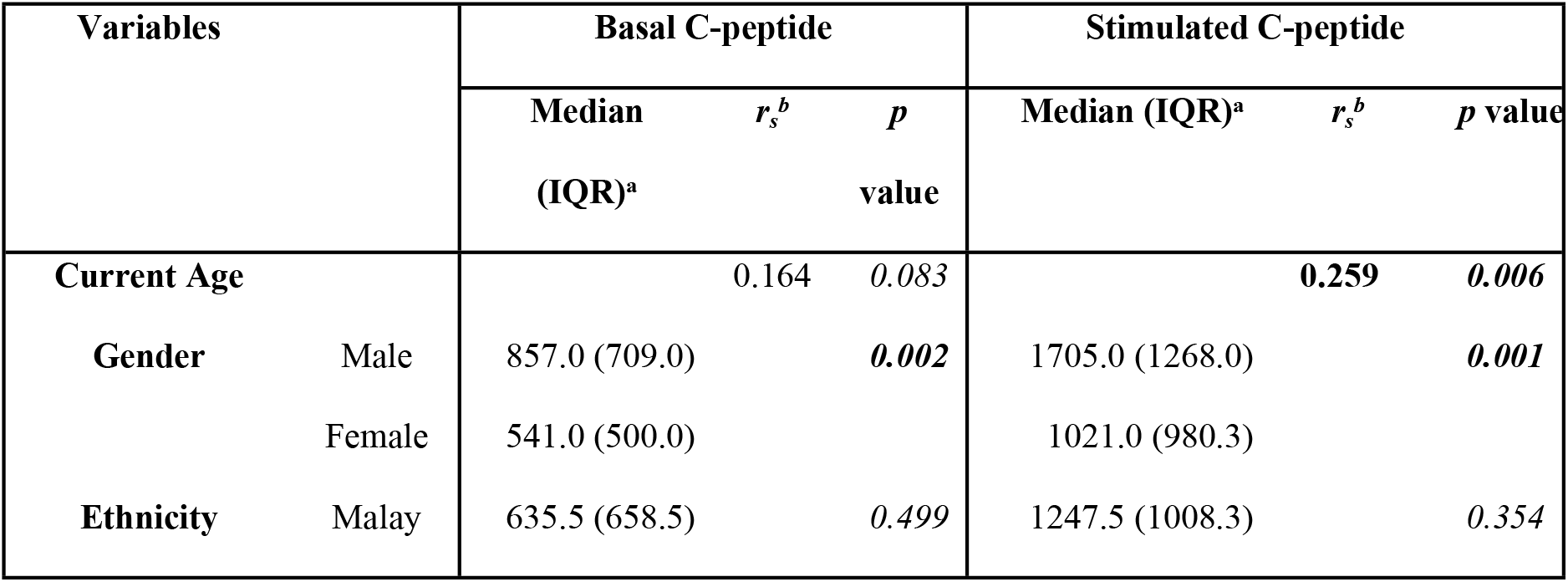
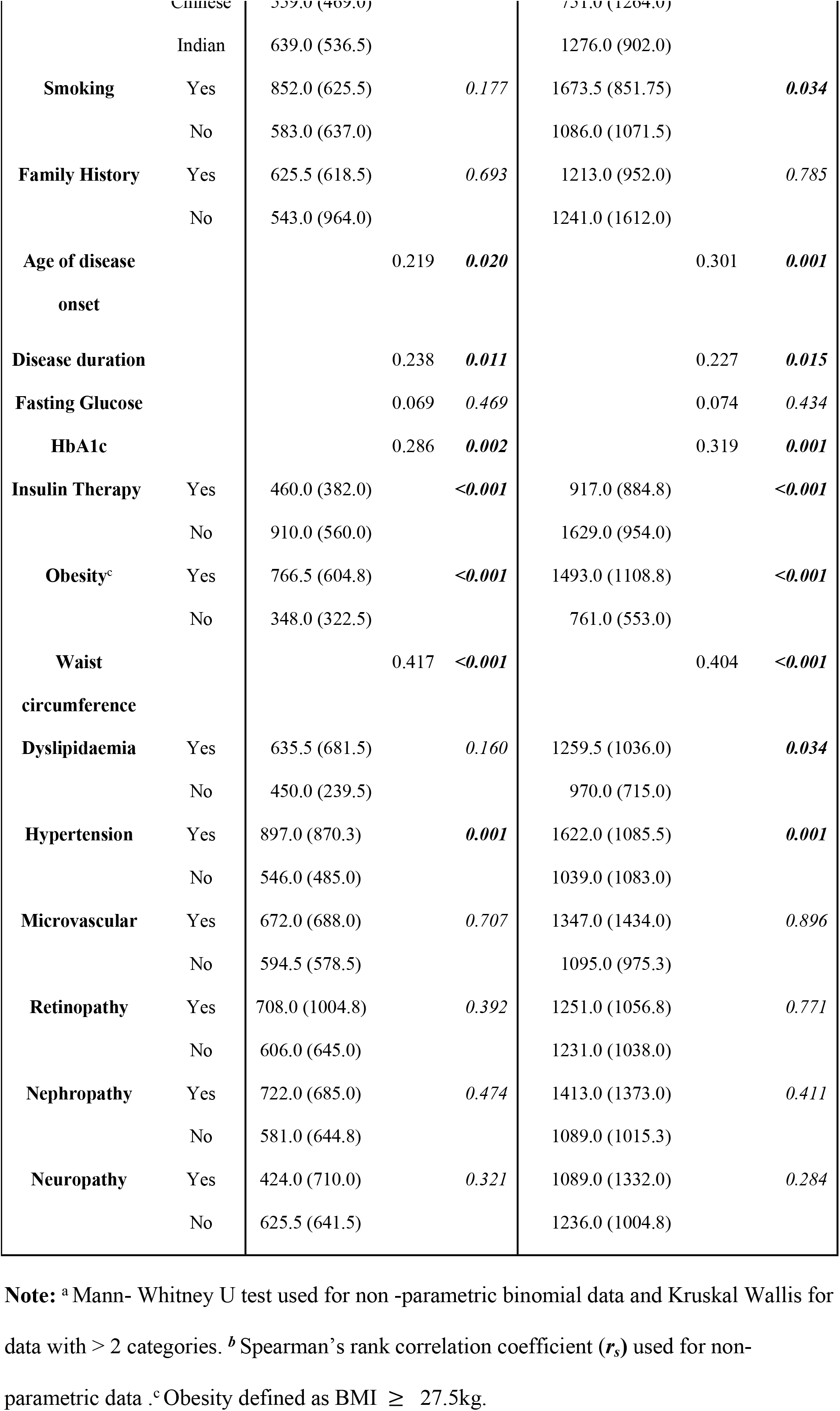
Bivariate analysis between C-peptide and clinical features, metabolic parameters and diabetes related complications.

**Table 4:** Factors independently associated with basal C-peptide levels from multivariable linear regression.

| Variables | Coefficient | 95% Confidence Interval |  | p value |
| --- | --- | --- | --- | --- |
|  |  | Lower Limit | Upper Limit |  |
| Insulin Therapy | - 392.8 | - 550.1 | - 235.4 | **<0.001** |
| Hypertension | 192.0 | 22.6 | 361.5 | **0.027** |
| Obesity | 252.6 | 78.9 | 426.3 | **0.005** |
| Female | - 266.8 | - 434.9 | - 99.6 | **0.002** |
| Nephropathy | 186.0 | 27.2 | 344.9 | **0.022** |
| Smoker | - 264.7 | - 501.5 | - 28.0 | **0.029** |

**Note:** Stepwise multiple linear regression was applied. The Durbin -Watson statistic, d=1.945 was between 1.5 and 2.5 and therefore data was not autocorrelated. Multicollinearity and homoscedasticity were checked and not found. A significant regression equation was found [F (6,104) = 13.076, p<0.001], with R<sup>2</sup> of 0.43.

**Table 5:** Factors independently associated with stimulated C-peptide levels from multivariable linear regression.

| Variable | Coefficient | 95% Confidence Interval |  | p value |
| --- | --- | --- | --- | --- |
|  |  | Lower Limit | Upper Limit |  |
| Insulin Therapy | - 393.4 | - 689.6 | - 97.3 | ***0.010*** |
| Hypertension | 533.5 | 201.6 | 865.4 | ***0.002*** |
| Obesity | 382.4 | 49.2 | 715.8 | ***0.025*** |
| Female | - 355.1 | - 644.1 | - 66.1 | ***0.017*** |
| Duration of Disease | - 7.2 | - 14.4 | - 0.2 | ***0.044*** |

**Note:** Stepwise multiple linear regression was applied. The Durbin -Watson statistic, $d=1.907$ was between 1.5 and 2.5 and therefore data was not autocorrelated. Multicollinearity and homoscedasticity were checked and not found. A significant regression equation was found [ $F(5,105) = 11.879$ , $p < 0.001$ ], with $R^2$ of 0.3.

## Discussion

We found our young onset T2DM population to have a predominance of females at 62% with strong family history of diabetes. Almost three quarter of them were obese by BMI. More than 90% had abdominal obesity associated with dyslipidaemia while almost one third had hypertension. This high prevalence of metabolic syndrome with multiple cardiovascular risk factors despite a young age and short disease duration is similar to the clinical characteristics reported in the literature (1,11,12). The median HbA1c of 8.5% (69 mmol/mol) in our study cohort indicated a relatively poorer glycaemic control compared to the HbA1c of 7.9% (63mmol/mol) for the general diabetic population in the country (13). Our findings concur with several studies done in the Asian region showing poorer glycaemic control in the young onset compared to the usual onset T2DM (12,14). The high percentage of insulin use within six months of diagnosis coupled with a low rate of subsequent weaning of insulin therapy in this study indicate a delayed diagnosis with high prevalence of severe hyperglycaemia at presentation requiring insulin followed by difficulties in restoring euglycemia despite intensive use of insulin. This is consistent with the high percentage of undiagnosed diabetes among the young age group of age 18 to 39 years at 77% from the National Health and Morbidity Survey in 2019 (15). On the contrary, only 13% were initiated on insulin at diagnosis and nearly all were weaned off insulin during the run-in period of two to six months using metformin titration and diabetes education in the Treatment Options for Type 2 Diabetes in Adolescents and Youth (TODAY) cohort (16). In the Asian region, Pan et al showed only 18% use of insulin in newly diagnosed young onset diabetes during the first year of disease despite the study population comprising approximately 10% of classical type 1 diabetes (14). There was a high prevalence of microvascular complication at 38% in our study cohort despite only after a median disease duration of two years. There was a striking predominance of DKD among the microvascular complications, with majority being microalbuminuria. Literature shows the prevalence of microalbuminuria is higher among individuals with young onset T2DM at diagnosis and tends to occur after a shorter duration of disease (2). The TODAY cohort showed a 6% prevalence of microalbuminuria at baseline but rose to17% after four years (16). The Pima Indian youth with T2DM (aged <20 years) had a much higher prevalence of microalbuminuria at diagnosis at 22% and was projected to reach 60% before the age of 30 (2). Kim et al also reported a high prevalence of microalbuminuria at 34.3%, nearly 2 folds higher in those with newly diagnosed young onset T2DM compared to those diagnosed after age 40 (11). This indicates an increased risk of early onset DKD with rapid progression among the young onset T2DM individuals compared to usual onset with possible ethnic variation.

The median C-peptide in our cohort was 619 pmol/L (basal) and 1231 pmol/L (glucagon stimulated). Based on the recommended thresholds of 250 pmol/L (basal) and 600 pmol/L (stimulated) C-peptide levels, majority (78-86%) of our patients have adequate beta-cell function (10). These results are comparable to other cohorts reported in the region (11,14). Kim et al reported a median basal C-peptide of 788 pmol/L and 2 hours post prandial C-peptide of 2023 pmol/L in their newly diagnosed young T2DM (11). Pan et al reported a mean basal C-peptide of 700 pmol/l with almost identical percentage of adequate beta cell function, 80-83% assessed using basal and glucagon stimulation C-peptide testing in their newly diagnosed young onset diabetes individuals consisting mainly of T2DM (14). This is in contrast to some other studies that reported significant beta cell dysfunction in newly diagnosed young onset T2DM (17,18). Even within the subgroup of insulin users, there was a high proportion of at least 70% having adequate beta cell function. Pan et al showed a lower percentage, approximately 50% of their young newly diagnosed DM on insulin with adequate basal and stimulated beta cell function using similar C-peptide cut off levels. However, their study population included those with T1DM (14). We postulated that the high fasting and stimulated C-peptide level shown in our study may be a surrogate marker of insulin resistance in this population (4,19). Therefore, we proceeded with post hoc-analysis to calculate modified HOMA-IR levels to assess insulin resistance in this cohort using basal C - peptide levels (20). We found a mean ± standard deviation (SD) modified HOMA-IR of 3.48±1.36. The modified HOMA-IR values showed a strong positive correlation with basal C-peptide levels, r*s=*0.90, p <0.001. Based on previous studies done in the Asian region, a HOMA-IR value of more than 2.5 has been used to define insulin resistance. (21,22). Using this cut off, 78% of our study population were found to be insulin resistant. This further supports our hypothesis that our recently diagnosed young onset T2DM population are predominantly insulin resistant as opposed to having inadequate beta cell function.

Multivariate analysis in this study revealed females to have lower basal and stimulated C-peptide levels than males. This may indicate a possible difference in beta-cell function between genders among the young onset T2DM. Despite the known preponderance of females in the young onset T2DM, there has been no conclusive evidence to suggest difference in beta-cell function between genders to date. Obesity and hypertension were independently associated with higher basal and stimulated C-peptide levels. This is presumably because an elevated C-peptide is known to be a surrogate marker of insulin resistance in individuals with metabolic syndrome phenotype (4). As expected, patients on insulin therapy were found to have significantly lower basal and stimulated C-peptide levels. There has been concern of suppression of endogenous insulin and C-peptide secretion in the literature in those on exogenous insulin therapy when beta cell function is assessed. However, it has been proven that acute normalization of glucose concentration or hypoglycaemia instead of the direct effect of the exogenous insulin is responsible for suppression of C-peptide levels during testing (4,23). Gjessing et al. has shown contrary to the hypothesis of beta cell function attenuation due to glucotoxicity, fasting hyperglycaemia in fact potentiates stimulated C-peptide levels during mixed meal tolerance test or glucagon stimulation test in T2DM (24). In this study, we ensured blood glucose of study participants was between 4.0 to 13.9 mmol/L at the time of testing to minimize the possible effect of extreme glucose concentrations on C-peptide values (5). We also found active smokers to have lower basal C-peptide levels than non-smokers. Although there are epidemiological studies confirming smoking as a risk factor for development of T2DM, the effect of smoking on beta cell function and insulin resistance is not conclusive (25–28). DKD was found to be independently associated with elevated basal C-peptide levels in this study. Previous studies done in the usual onset T2DM with longer disease duration showed an opposite relationship with lower C-peptide levels associated with DKD (5,19). In our study cohort, there was a high prevalence of DKD early in the disease despite majority of them having adequate beta-cell function. Beta cell dysfunction may therefore not be the main driving force behind the early development of DKD in this cohort. The high prevalence of metabolic syndrome phenotype, elevated serum C-peptide and modified HOMA-IR levels may indicate insulin resistance as the key factor contributing to the early onset DKD. A large Scandinavian data-driven cluster analysis has identified a subgroup of diabetes labelled as Severe Insulin Resistance Diabetes (SIRD) characterized by severe insulin resistance and high BMI. The risk of DKD was significantly increased in this cluster of patients independent of their glycaemic control illustrating the relationship between insulin resistance and kidney disease. Insulin resistance has been associated with increased salt sensitivity, glomerular hypertension and hyperfiltration resulting in deteriorating in kidney function in these patients (29).

This study had a few limitations. The small sample size may have underscored the power in detecting the association between C-peptide with metabolic parameters and diabetes related complications. The cross-sectional study design was not able to establish causal relationship between measured variables and outcomes. Our study population comprised of patients attending diabetes clinic in two urban tertiary hospital was not fully representative of the general population of young onset T2DM in the country. This is the first study assessing beta cell function in recently diagnosed, exclusively young onset T2DM in the country and one of the few in Asia. Various measures to minimize effect of confounding factors on C-peptide measurement were undertaken including stringent eligibility criteria, exclusion of extreme blood glucose levels and omission of exogenous insulin on the day of testing to improve the accuracy of C-peptide testing.

## Conclusion

Most of our recently diagnosed young onset T2DM individuals have adequate beta cell function. There was a high rate of metabolic syndrome in this study cohort associated with early onset DKD. The glycaemic control was poor despite high rates of insulin use. Elevated C-peptide levels associated with obesity, hypertension and DKD suggest that insulin resistance rather than beta cell dysfunction as the key driving factor of complications. Our study paves the direction for future prospective studies involving larger cohort of individuals with direct measurement of insulin sensitivity to confirm our findings. Treatment strategies in this population should be tailored, focussing on early diagnosis before development of severe hyperglycaemia with a combination of intensive lifestyle modification and pharmacotherapy to address obesity, insulin resistance and avoiding overtreatment with insulin. These individuals should be screened for microvascular complications especially DKD upon diagnosis and monitored closely thereafter to prevent premature morbidity and mortality.

## Data Availability

All relevant data are within the manuscript and its supporting information files

